# Development and application of an algorithm for statin-induced myopathy based on electronic health record-derived structured elements

**DOI:** 10.1101/2023.04.24.23289059

**Authors:** Akinyemi Oni-Orisan, Meng Lu, Jonathan A. Peng, Ronald M. Krauss, Carlos Iribarren, Marisa W. Medina

**Affiliations:** Department of Clinical Pharmacy, Institute for Human Genetics, University of California San Francisco, San Francisco CA 94143, USA; Kaiser Permanente Division of Research, 2000 Broadway, Oakland, CA 94612, USA; Department of Cardiology, Kaiser Permanente, Santa Rosa, CA 95403, USA; Department of Medicine, Department of Pediatrics, University of California San Francisco, Oakland CA 94609, USA; Department of Pediatrics, University of California San Francisco, Oakland CA 94609, USA

**Author notes:** co-corresponding authors, Correspondence to: Marisa W. Medina, 5700 Martin Luther King Jr. Way, Oakland, CA 94609, Carlos Iribarren 2000 Broadway, Oakland, CA 94612. All authors have seen and approve the manuscript. **Competing Interests:** None.

**Keywords:** Adverse drug reaction, creatine kinase, EHR, myotoxicity, real world evidence

## Abstract

**Objective:** Considering the non-specific nature of muscle symptoms, studies of statin-induced myopathy (SIM) in electronic health records require accurate algortihms that can reliably identify true statin-related cases. However, prior algorithms have been constructed in study populations that preclude broad applicability. Here we developed and validated an algorithm that accurately defines SIM from electronic health records using structured data elements and conducted a study of determinants of SIM after applying the algorithm.

**Materials and Methods:** We used electronic records from an integrated health care delivery system (including comprehensive pharmacy dispensing records) and defined SIM as elevated creatine kinase (CK) ≥4 x upper limit of normal. A diverse cohort of participants receiving a variety of statin regimens met the criteria for study inclusion.

**Results:** We identified multiple conditions strongly associated with elevated CK independent of statin use. A 2-step algorithm was developed using these all-cause conditions as secondary causes (step 1) along with evidence of a statin regimen change (step 2). We identified 1,262 algorithm-derived statin-induced elevated CK cases. Gold standard SIM cases determined from manual chart reviews on a random subset of the all-cause elevated CK cases were used to validate the algorithm, which had a 76% sensitivity and 77% specificity for detecting the most certain cases. Pravastatin use was associated with a 2.18 odds (95% confidence interval 1.39-3.40, P=0.0007) for statin-induced CK elevation compared to lovastatin use after adjusting for dose and other factors.

**Conclusions:** We have produced an efficient, easy-to-apply methodological tool that can improve the quality of future research on statin-induced myopathy.

## BACKGROUND and SIGNIFICANCE

Statin therapy reduces morbidity and mortality associated with atherosclerotic cardiovascular disease (ASCVD). Specifically, results from randomized controlled trials estimate that for every ∼40 mg/dL lowering of low-density lipoprotein cholesterol (LDL-C) by statin treatment, the risk of an ASCVD event is reduced by 21% and the risk of all-cause death is reduced by 12%^1^. Consequently, national guidelines recommend moderate-to high-intensity statin therapy first-line for the primary and secondary prevention of ASCVD^2^. This endorsement has led to high prescribing rates of intensive statin therapy in the United States^3^.

Despite the aforementioned well-documented efficacy and increased prescribing of statins, ASCVD remains the leading cause of mortality in the United States and globally. More widespread statin use, which would translate to improved cardiovascular outcomes on a population-wide scale, is hampered by patient non-adherence^4^. The 1-year discontinuation rate for statin initiators is as high as 70% in some populations^5^. Importantly, statin-associated muscle symptoms (SAMS) are a major cause of statin discontinuation^6^ with reported rates as high as ∼25% in multiple observational studies^7–9^.

Further investigation of statin myotoxicity can provide guidance on detecting, avoiding, and managing SAMS. Among the broad spectrum of SAMS, statin-induced myopathy (SIM) is defined in part by elevated blood creatine kinase (CK) levels^10^. Unlike self-reported muscle symptoms, which are often documented through free text in the electronic health records, this objective measure allows for efficient detection of SIM in the records of large population-based cohorts where manual chart review is not feasible and natural language processing techniques are prohibitively expensive^11^. However, CK is a non-specific biomarker for muscle injury. We need robust algorithms that can correctly differentiate myopathy caused by statins versus other causes. The quality of these algorithms has implications for genetic and epidemiologic studies that rely on identifying true SIM cases for producing accurate results. Prior algorithms for this purpose have been constructed in small or homogenous populations and may not have broad applicability^12,13^.

## OBJECTIVE

The objective of this study was to characterize SIM in a large, diverse population of patients during routine clinical care. We first identified conditions significantly associated (in a clinically meaningful way) with all-cause elevated CK unrelated to statin therapy. We used these conditions as exclusionary factors to develop and validate an algorithm designed to extract true SIM cases from electronic health records. Finally, we used these true cases to identify clinical factors that increase patient risk of SIM.

The findings from this study have produced a methodological tool that can improve the quality of future research on SIM and aid clinicians in avoiding this adverse effect.

## MATERIALS and METHODS

### Data source

All participants were members of Kaiser Permanente Northern California (KPNC), an integrated health care delivery system featuring all-inclusive services (e.g., inpatient, outpatient, pharmacy, laboratory) with the exception of dental care and vision. KPNC electronic health records containing demographics, laboratory records, pharmacy records, and International Classification of Diseases (ICD) diagnostic codes (ICD-9 and ICD-10) between the years 2000 and 2018 were used. Laboratory records included CK and fasting total cholesterol, LDL-C, high-density lipoprotein cholesterol (HDL-C), triglycerides, glucose, and hemoglobin A1c levels. Pharmacy data included dispensing records for simvastatin, lovastatin, atorvastatin, pravastatin and rosuvastatin. Diagnoses and diagnostic codes included toxic myopathy, rhabdomyolysis, hypothyroidism, hypertension, gout, diabetes, opioid abuse, skeletal disease, human immunodeficiency virus/acquired immunodeficiency syndrome, acute myocardial infarction, major surgery, acute kidney failure (or dialysis), accidents/injuries, falls, sepsis/septic shock, coma, stroke, convulsions, paraplegia, dehydration, pneumonia, and overexertion from strenuous movement/load (codes are shown in **Supplementary Table 1**). Other electronic health records included for this study were body mass index (BMI), systolic blood pressure (SBP), and diastolic blood pressure (DBP). The study was approved by the Kaiser Foundation Research Institute and University of California San Francisco Institutional Review Boards.

### Study population

Statin users were defined as initiators of simvastatin, lovastatin, atorvastatin, pravastatin or rosuvastatin monotherapy at age 45-70 with evidence of “persistent statin use” defined by the interval from statin initiation until elevated CK or the end of the followup period (December 31, 2018). Pharmacy dispensing records were used to estimate a mean possession ratio for each statin user as the measure of adherence (cumulative days’ supply dispensed within follow-up period ÷ total days of follow-up). We used fold change from the upper limit of normal (ULN) as the unit for CK and defined elevated CK at ≥4 x ULN (336 IU/L in males and 176 IU/L in females)^14^. Statin nonusers were defined as individuals with no records of statin dispensing.

A statin user met the criteria for a SIM case if they had (1) ≥1 incident of all-cause elevated CK within the maximum follow-up window following statin initiation and (2) no elevated all-cause elevated CK within the 5 years prior to statin initiation. If a statin user case had >1 record of elevated CK within the maximum follow-up window following statin initiation, the earliest level was chosen as the index CK. The statin record with a day’s supply window overlapping the index CK was considered the index pharmacy record. If multiple statins within the study time window for a participant overlapped with the index CK, the dispensing record most immediately preceding the elevated CK temporally, was considered the index pharmacy record. All statin users meeting the criteria for a case of all-cause elevated CK were included in the analysis. A statin user met the criteria for a control if they had ≥1 CK level <2 x ULN, but no CK levels ≥2 x ULN within the maximum follow-up time frame of the study. If a statin user control had >1 CK level within the time frame of the study, the earliest CK was chosen as the index CK with the date of the index CK designated as the index date. Among statin users meeting the criteria for a control, participants were selected randomly for inclusion in the analysis until a 4:1 control-to-case ratio (matched by age at statin initiation, sex, and race/ethnicity) was reached.

To facilitate the identification of factors associated with CK elevation, we also defined a cohort of statin nonuser cases (≥1 incident of all-cause elevated CK in patients with no records of statin dispensing). Statin cases were matched in a 1:1 ratio with the statin nonuser cases by sex, race/ethnicity and age of elevated CK. Finally, a set of statin nonuser controls (≥1 CK level <2 x ULN, but no CK levels ≥2 x ULN) were matched to the statin nonuser cases in a 2:1 ratio by sex, race/ethnicity, and age of statin initiation.

### Identification of factors associated with all-cause elevated CK and algorithm-predicted statin-induced elevated CK

The independent association between acute conditions known to be associated with CK elevations (AMI, major surgery, acute kidney failure/dialysis, accidents/injuries, falls, sepsis/septic shock, coma, stroke, convulsions, paraplegia, dehydration, pneumonia, overexertion, uncontrolled diabetes, uncontrolled hypothyroidism, hypertensive urgency, uncontrolled gout)^13,15–17^ and all-cause elevated CK among statin cases versus controls was determined. For this analysis, acute conditions were considered to be linked to an elevated CK if the diagnosis code for the condition occurred within 7 days of that elevated CK. Rare covariates (prevalence <0.1%) were not included in the model. We defined uncontrolled gout as uric acid >7.2 mg/dL in males and >6.5 mg/dL in females, hypothyroidism as thyroid stimulating hormone >3mIU/L, uncontrolled diabetes as HbA1C >9%, and hypertensive urgency as SBP>180 mmHg and/or DBP>110 mmHg. Identical analyses were used in statin nonusers to determine the association between acute conditions and all-cause elevated CK among statin nonuser cases versus controls. An interaction test was conducted to compare the odds ratios (ORs) of all-cause elevated CK for each acute condition between statin users and nonusers. Acute conditions associated with all-cause elevated CK in statin nonusers were used in the development of an algorithm (as described in greater detail below) to identify CK levels possibly unrelated to statin therapy. Predictors of statin-induced elevated CK were then determined among algorithm-predicted statin-induced CK elevation cases and matching controls (matched by age of statin initiation, sex and race/ethnicity) with a ratio of 1 case: 2 controls. Additional covariates added to this model included statin type, statin daily milligram strength, and statin defined daily dose units, as previously described^18^. Briefly, the statin defined daily dose accounts for differences in LDL-C lowering potency among statin types^18^. We set 1.0 defined daily dose units equal to 40 mg of lovastatin daily and all of its equipotent counterparts. Relative LDL-C lowering potency among statin types was based on the revised equivalency chart^18^.

### Algorithm development

Step 1: Among the full set of all-cause elevated CK statin user cases (as defined above), participants were carried forward to step 2 if their index elevated CK level date was outside of a 7-day window following any acute condition previously found to be associated with all-cause elevated CK in statin nonusers (see section above)

Step 2: Among participants carried forward to step 2, those with evidence of statin discontinuation (no subsequent statin dispensing record within 60 days following the day’s supply of the index pharmacy record) were kept.

The index CK levels among participants kept in Step 2 were deemed to be attributed to true cases of SIM. The index CK levels of all other participants were considered to be elevated due to reasons other than statin use.

### Algorithm validation

The electronic charts of 99 participants among the all-cause elevated CK cases were randomly selected for manual chart review by two independent reviewers (AO and JP) to determine the likelihood of statin use as a causative factor for the elevated CK. Assessment of algorithm performance requires distribution across all possible test outcomes (e.g., examples of false positives, false negatives, true positives, and true negatives). To increase the chance of fairly uniform distribution, the 99 participants for chart review were randomly chosen such that 33 were chosen from the subset of all-cause elevated CK cases that did not make it past step 1 of the algorithm, 33 were chosen from the subset that made it through step 1 but not step 2, and 33 were chosen from the subset that made it through steps 1 and 2. Each of the 99 charts were manually reviewed and classified into one of four categories describing how likely the index elevated CK was statin-induced versus another cause (Certain, Probable, Possible, and Unlikely) based on the modified version of a prior classification system (**Supplementary Table 2**)^12^. Inconsistencies between reviewers were resolved by a final joint review to reach consensus. Gold standard statin-induced elevated CK cases were defined in three different ways (i. Certain, ii. Certain + Probable, iii.

Certain + Probable + Possible) and were used to measure the performance of the algorithm. Sensitivity, specificity, positive predictive value (PPV), and negative predictive value (NPV) were calculated as performance metrics of the algorithm.

### Statistical analyses

All univariate case-control comparisons of baseline characteristics were done using t-tests for continuous variables that were normally distributed (all but triglycerides), the Kruskal Wallis test for continuous variables that were not-normally distributed (i.e., triglycerides) and Chi-square tests for categorical variables. The relationship between factors and all-cause elevated CK or <u>algorithm-predicted statin-induced elevated CK</u>was estimated using conditional logistic regression first univariately and then in the multivariable context

## RESULTS

### Descriptive characteristics

We identified 998,054 eligible statin users in the electronic health records. Among these, 370,859 had measured CK of whom 5,430 met the definition of a statin user case. As comparators, we identified three age, sex and race/ethnicity matched cohorts: i) statin user controls (n=13,171), ii) statin nonuser cases (n=5,726), iii) statin nonuser controls (n=22,904). In the statin user cases, the median time to CK elevation from initiation was 66.0 ± 49.0 months (mean±standard deviation [SD]) with a mean possession ration of 0.77. As a positive control, we confirmed that both the statin user and statin nonuser cases had significantly higher frequencies of clinically diagnosed toxic myopathy or rhabdomyolysis as compared to their respective control cohorts. Demographics and clinical characteristics of all four cohorts are described in **Table 1**.

**Table 1.** Demographics and clinical characteristics of statin users and statin non-users at all cause CK elevation

|  | <u>At First Statin Prescription</u> |  | <u>At Time of Elevated CK</u> |  |
| --- | --- | --- | --- | --- |
|  | **Elevated CK CASES** | **CONTROLS** | **Elevated CK CASES** | **CONTROLS** |
| | Statin users with CK $\geq 4$ ULN†<br>n= 5,430 | Statin Users with CK always < 2 ULN<br>n=13,171 | Statin users with CK $\geq 4$ ULN†<br>n= 5,430 | Statin Users with CK always < 2 ULN<br>n=13,171 |
| **Sociodemographic Characteristics** |  |  |  |  |
| Age, years, mean ± SD | 62.6 ± 12.5 | 62.5 ± 11.9 | 62.6 ± 12.5 | 62.6 ± 12.5 |
| Gender, n (%) |  |  |  |  |
| Male | 2,486 (45.8%) | 6,011 (45.6%) | 2,608 (45.5%) | 10,432 (45.5%) |
| Female | 2,944 (54.2%) | 7,160 (54.4%) | 3,118 (54.5%) | 12,472 (54.5%) |
| Race\Ethnicity, n (%) |  |  |  |  |
| White | 2,969 (54.7%) | 7,208 (54.7%) | 3,122 (54.5%) | 12,488 (54.5%) |
| Black | 659 (12.1%) | 1,507 (11.4%) | 694 (12.1%) | 2,776 (12.1%) |
| Hispanic | 620 (11.4%) | 1,465 (11.1%) | 659 (11.5%) | 2,636 (11.5%) |
| Asian | 747 (13.8%) | 1,917 (14.6%) | 799 (14.0%) | 3,222 (14.1%) |
| Other\Missing | 435 (8.0%) | 1,074 (8.2%) | 452 (7.9%) | 1,782 (7.8%) |
| **Cardiometabolic Factors at the Time of CK Elevation\*, (n), mean ± SD** |  |  |  |  |
| BMI, Kg/m<sup>2</sup> | (5,060), 29.8 ± 6.7 | (11,985), 29.4 ± 6.0 | (5,349), 29.8 ± 6.8 | (19,167), 29.1 ± 6.1 |
| Total cholesterol, mg/dL | (5,389), 170.2 ± 45.0 | (13,093), 176.2 ± 40.5 | (5,680), 169.9 ± 45.1 | (20,833), 179.8 ± 44.0 |
| LDL cholesterol, mg/dL | (5,393), 92.2 ± 35.5 | (13,090), 96.5 ± 32.3 | (5,686), 91.8 ± 35.4 | (20,880), 100.3 ± 35.9 |
| HDL cholesterol, mg/dL | (5,385), 48.1 ± 14.8 | (13,080), 50.8 ± 14.1 | (5,676), 48.2 ± 14.9 | (20,806), 50.5 ± 14.2 |
| Triglycerides, mg/dL | (5,361), 157.7 ± 141.6 | (13,057), 150.1 ± 117.5 | (5,648), 157.7 ± 155.6 | (20,703), 150.5 ± 115.7 |
| Fasting glucose, mg/dL | (5,034), 123.0 ± 53.2 | (12,487), 116.9 ± 41.1 | (5,286), 122.9 ± 52.8 | (19,467), 117.5 ± 42.8 |
| HbA1c, % | (3,955), 6.8 ± 1.5 | (8,422), 6.6 ± 1.3 | (4,203), 6.8 ± 1.5 | (13,481), 6.6 ± 1.4 |
| SBP, mmHg | (3,998), 128.4 ± 21.8 | (8,817), 128.0 ± 15.7 | (4,274), 128.6 ± 21.8 | (14,482), 128.2 ± 16.8 |
| DBP, mmHg | (3,998), 69.3 ± 13.6 | (8,817), 71.6 ± 10.4 | (4,274), 69.3 ± 13.6 | (14,482), 71.8 ± 11.0 |
| Uric acid, mg/dL | (1,743), 6.9 ± 2.1 | (3,578), 6.4 ± 1.8 | (1,831), 6.9 ± 2.1 | (5,686), 6.4 ± 1.9 |
| **Statin Type** |  |  |  |  |
| Atorvastatin | 284 (5.2%) | 552 (4.2%) | 906 (16.7%) | 1,524 (11.6%) |
| Lovastatin | 3,900 (71.8%) | 9,511 (72.2%) | 2,545 (46.9%) | 7,038 (53.4%) |
| Pravastatin | 65 (1.2%) | 167 (1.3%) | 167 (3.1%) | 329 (2.5%) |
| Rosuvastatin | 6 (0.1%) | 8 (0.1%) | 15 (0.3%) | 27 (0.2%) |
| Simvastatin | 1,175 (21.6%) | 2,933 (22.3%) | 1,797 (33.1%) | 4,253 (32.3%) |
| **Time between fist prescription and elevated CK in cases or index CK elevation date for controls, months, mean ± SD** |  |  |  |  |
| | 66.0 ± 49.0 | 57.5 ± 44.0 | | |
| **% days covered by statins between first prescription and SIM in cases or index date in controls (%)** |  |  |  |  |
| | | | 76.9 ± 22.3 | 81.9 ± 18.3 |

\*single measure preceding closest in time.

In the statin users, the cases had higher BMI, lower LDL-C and HDL-C, and higher triglycerides and fasting glucose compared to statin user controls. Nearly identical differences were observed in the statin nonuser cohort. The most common statin prescription among statin users was lovastatin, thus lovastatin was considered the reference for analyses of statin type. Statin type and dose characteristics are shown in **Table 2, Supplementary Table 3**.

**Table 2.** Performance of algorithms for SIM detection based on manual chart review

| **SIM Case definition** | **Algorithm defined cases** | **Positive Chart review cases** | **Negative chart review cases** | **Sensitivity** | **Specificity** |
| --- | --- | --- | --- | --- | --- |
| **Certain only** | Positive | 16 | 17 | 76% | 77% |
|  | Negative | 5 | 57 |  |  |
| **Certain + Probable** | Positive | 19 | 14 | 66% | 79% |
|  | Negative | 10 | 52 |  |  |
| **Certain + Probable + Possible** | Positive | 25 | 8 | 58% | 85% |
|  | Negative | 18 | 44 |  |  |

### Predictors of all-cause elevated CK among statin users

The majority of our prescpecified conditions were each independently associated with all-cause elevated CK in both the statin user and statin non-user cohorts (**Supplementary Table 4**). In a multivariate model, nearly all of the acute, and 6 of the 8 chronic conditions remained associated with elevated all-cause CK in the statin user cohort (**Supplementary Table 5**). The most common conditions linked to all-cause elevated CK were MI/STEMI/MSTEMI/RP (25.76%), acute kidney failure (20.52%), and sepsis/septic shock (13.95%).

### Algorithm to predict cases of statin-induced elevated CK among all-cause elevated CK

There were a total of 5,430 statin users that met study criteria as a case of all-cause elevated CK. Based on the clear association between 16 acute conditions and all-cause CK irrespective of statin use, we developed an algorithm to identify true cases of SIM. Following Step 1 of the algorithm, 1,942 participants remained (**Figure 1**). The final step of the algorithm (Step 2) yielded a total of 1,262 participants deemed to have true SIM. Among the algorithm-predicted cases of statin-induced elevated CK, participants were 58.9 ± 13.0 (mean ± SD) years of age at the time of statin therapy initiation with a median time of elevated CK occurring 62.0 ± 48.5 (mean ± SD) days following statin initiation.

**Figure 1:**
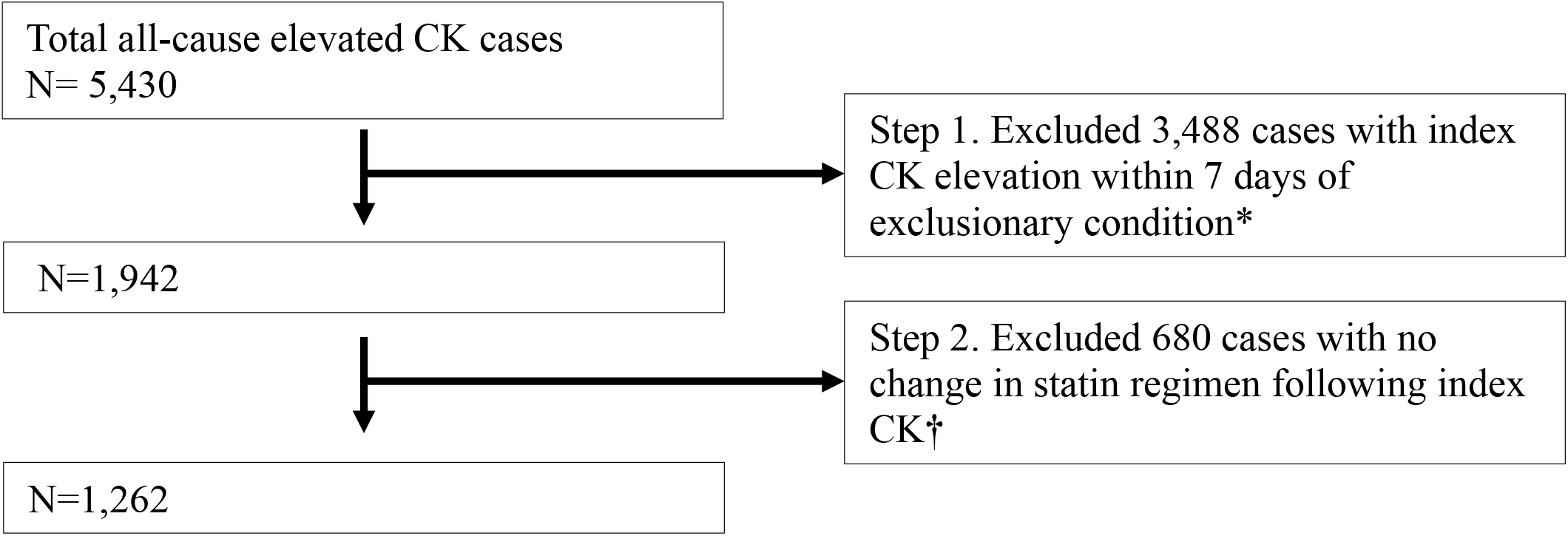
Algorithm to derive statin-induced CK cases. *Flow chart describing numbers carried forward at each step to narrow from all-cause elevated CK cases to statin-induced elevated CK cases* *\*List of acute conditions we deemed in our logistic regression to be causative of all-cause CK elevation* *†Evidence of statin regimen change included: 1) continuation at same type/dose for ONLY one refill, 2) lowered dose (of same type) or different type following elevated CK, or 3) no further statin refills*

### Algorithm performance and validation

Of the electronic charts from the 99 all-cause elevated CK cases selected for manual chart review, the likelihood of SIM was determined to be Certain for 21, Probable for 8, Possible for 14, and Unlikely for 52. Charts were unavailable for review from 4 individuals.

Each step of the algorithm resulted in additive improvement for specificity and PPV metrics based on a strong ability to correctly identify true negatives (**Table 2, Supplemental Table 6**). However, as expected, each step of the algorithm also resulted in reduced sensitivity due to false negatives. When the most liberal approach for gold standard statin-induced elevated CK cases was selected (Certain + Probable + Possible), Step 1 alone of the algorithm improved specificity from 0% to 58%, and reduced sensitivity from 100% to 93%. Using the same gold standard, implementation of the full algorithm (Steps 1 + 2) resulted in improved specificity (85% from 58%)and reduced sensitivity (from 93% to 58%) compared to step 1 alone.

### Description of algorithm-determined statin-induced CK elevation

The full algorithm (Steps 1 + 2) was applied to the full cohort of statin users with all-cause CK elevations to extract statin-induced CK cases (n=1,257). Sociodemographic and statin prescription characteristics are described in **Table 3**.

**Table 3.** Sociodemographic and statin prescription characteristics of the SIM Case and Control cohorts.

|  |  | <u>At First Statin Prescription</u> |  |  | <u>At time of elevated or index CK</u> |  |  |
| --- | --- | --- | --- | --- | --- | --- | --- |
|  |  | SIM Cases<br>n= 1,257 | Controls<br>n=3,047 | P | SIM Cases<br>n= 1,257 | Controls<br>n=3,047 | P |
| **Age, years, mean ± SD** |  | 58.9 ± 13.0 | 59.1 ± 12.3 |  |  |  |  |
| **Gender, n (%)** | Male | 599 (47.7%) | 1,438<br>(47.2%) |  |  |  |  |
|  | Female | 658 (52.3%) | 1,609<br>(52.8%) |  |  |  |  |
| **Race\ethnicity, n (%)** | White | 616 (49.0%) | 1,507<br>(49.5%) |  |  |  |  |
|  | Black | 193 (15.4%) | 455 (14.9%) |  |  |  |  |
|  | Hispanic | 151 (12.0%) | 365 (12.0%) |  |  |  |  |
|  | Asian | 187 (14.9%) | 458 (15.0%) |  |  |  |  |
|  | Other\missing | 110 (8.8%) | 262 (8.6%) |  |  |  |  |
| **Statin type** | Atorvastatin | 80 (6.4%) | 141 (4.6%) | 0.02 | 199 (15.8%) | 361 (11.8%) | 0.0004 |
|  | Lovastatin | 843 (67.1%) | 2,198<br>(72.1%) | 0.0005 | 542 (43.1%) | 1,568<br>(51.5%) | <0.0001 |
|  | Pravastatin | 19 (1.5%) | 32 (1.1%) | 0.20 | 47 (3.7%) | 67 (2.2%) | 0.004 |
|  | Simvastatin | 315 (25.1%) | 676 (22.2%) | 0.04 | 469 (37.3%) | 1,051<br>(34.5%) | 0.08 |
| **Daily dose by statin type** | Atorvastatin <20 mg | 30 (2.4%) | 54 (1.8%) | 0.14 | 54 (4.3%) | 103 (3.4%) | 0.14 |
|  | 20 to <40 mg | 24 (1.9%) | 45 (1.5%) | 0.30 | 59 (4.7%) | 96 (3.2%) | 0.01 |
|  | 40 to <80 mg | 11 (0.9%) | 20 (0.7%) | 0.44 | 63 (5.0%) | 124 (4.1%) | 0.17 |
|  | ≥80 mg | 15 (1.2%) | 22 (0.7%) | 0.12 | 23 (1.8%) | 38 (1.2%) | 0.15 |
|  | Lovastatin <20 mg | 272 (21.6%) | 742 (24.4%) | 0.04 | 153 (12.2%) | 507 (16.6%) | 0.0002 |
|  | 20 to <40 mg | 296 (23.5%) | 803 (26.4%) | 0.04 | 179 (14.2%) | 564 (18.5%) | 0.0008 |
|  | 40 to <80 mg | 271 (21.6%) | 642 (21.1%) | 0.75 | 198 (15.8%) | 471 (15.5%) | 0.81 |
|  | ≥80 mg | 4 (0.3%) | 11 (0.4%) | 0.82 | 12 (1.0%) | 26 (0.9%) | 0.75 |
|  | Pravastatin <20 mg | 3 (0.2%) | 2 (0.1%) | 0.13 | 11 (0.9%) | 20 (0.7%) | 0.44 |
|  | 20 to <40 mg | 7 (0.6%) | 20 (0.7%) | 0.70 | 17 (1.4%) | 22 (0.7%) | 0.05 |
|  | 40 to <80 mg | 9 (0.7%) | 8 (0.3%) | 0.03 | 17 (1.4%) | 24 (0.8%) | 0.08 |
|  | ≥80 mg | 0 (0.0%) | 2 (0.1%) | 0.36 | 2 (0.2%) | 1 (0.0%) | 0.15 |
|  | Simvastatin <20 mg | 28 (2.2%) | 53 (1.7%) | 0.28 | 115 (9.1%) | 221 (7.3%) | 0.04 |
|  | 20 to <40 mg | 54 (4.3%) | 138 (4.5%) | 0.68 | 126 (10.0%) | 324 (10.6%) | 0.55 |
|  | 40 to <80 mg | 166 (13.2%) | 369 (12.1%) | 0.30 | 179 (14.2%) | 420 (13.8%) | 0.69 |
|  | ≥80 mg | 67 (5.3%) | 116 (3.8%) | 0.02 | 49 (3.9%) | 86 (2.8%) | 0.06 |
| **Defined daily dose** | ≤ ¼ (Low) | 282 (22.4%) | 764 (25.1%) |  | 80 (6.4%) | 262 (8.6%) |  |
|  | 1/4 - <1 (Low moderate) | 333 (26.5%) | 864 (28.4%) |  | 191 (15.2%) | 632 (20.7%) |  |
|  | 1 (Moderate) | 300 (23.9%) | 698 (22.9%) |  | 358 (28.5%) | 855 (28.1%) |  |
|  | 1 - < 3 (Moderate High) | 249 (19.8%) | 562 (18.4%) |  | 297 (23.6%) | 703 (23.1%) |  |
|  | >3 (High) | 93 (7.4%) | 159 (5.2%) |  | 331 (26.3%) | 595 (19.5%) |  |
| **Statin daily dose (irrespective of type)** | <20 mg | 333 (26.5%) | 851 (27.9%) |  | 124 (9.9%) | 387 (12.7%) |  |
|  | 20-40 mg | 381 (30.3%) | 1,006<br>(33.0%) |  | 268 (21.3%) | 836 (27.4%) |  |
|  | 40-80 mg | 457 (36.4%) | 1,039<br>(34.1%) |  | 577 (45.9%) | 1,275<br>(41.8%) |  |
|  | ≥80 mg | 86 (6.8%) | 151 (5.0%) |  | 288 (22.9%) | 549 (18.0%) |  |
| **Time between first prescription and SIM in cases or index date for controls (months), mean ± SD** |  | 62.0 ± 48.5 | 57.2 ± 44.6 | 0.002 |  |  |  |
| **% days covered by statins between first prescription and SIM in cases or index date in controls (%)** |  |  |  |  | 83.1 ± 24.8 | 86.7 ± 21.5 | <0.0001 |

Among the algorithm-predicted cases, 52% of patients were female and 49% white. The median age of statin initiation was 58.9±13.0 (mean±SD) years and the median time-to-elevated CK following initiation was 62.0±48.5 (mean±SD) months. The majority of patients were first prescribed lovastatin (67.1%) or simvastatin (25.1%) encompassing a wide range of doses from low to moderately high. Rosuvastatin users were omitted from the analysis due to the very small number of total users. Like the cases, the majority of algorithm-predicted statin-induced CK elevation controls were also first prescribed either lovastatin (72.1%) or simvastatin (22.2%). Algorithm-predicted statin-induced CK cases had reduced rates of statin adherence compared to matched controls (average 83.1% vs. 86.7% mean possession ratio covered; p<0.00001).

### Predictors of statin-induced CK elevation

Both statin type and statin dose were independently associated with SIM risk (**Table 4**). In particular, use of atorvastatin at first prescription (OR 1.49, 95% confidence interval [CI], 1.11-1.99, P=0.007) and at the time of CK elevation (OR 1.35, 95% CI 1.01-1.81, P=0.04) was associated with increased risk of statin-induced elevated CK compared to lovastatin reference (**Table 4**). Notably, pravastatin and simvastatin use at the time of elevated CK were each also associated with greater risk of SIM compared to lovastatin. In terms of statin daily dose, a step-wise dose response relationship was observed with increasing milligram strength at the time of elevated CK a strong predictor of statin-induced elevated CK. For example, at the time of elevated CK, statin users with a 40-79 mg daily dose had a 1.47 OR (95% CI 1.17-1.83, P=0.0007) compared to the lowest reference dose of <20mg daily dose (**Table 4**). When assessed as a continuous variable, statin daily dose (irrespective of type) at the time of elevated CK was highly associated with SIM (P=0.0004), **Supplementary Table 7**. In addition, as we also considered change in statin dose between the initial statin prescription and the prescription at the time of elevated CK. Compared to individuals who did not change their daily dose, those that increased their statin dose had a greater risk of SIM (OR 1.22, 95% CI 1.04-1.43), **Supplementary Table 8**.

**Table 4.** Independent associations of statin type, statin daily dose (irrespective of type) and comorbidities with SIM case/control status.

|  | <u>At time of first prescription</u> |  | <u>At time of elevated/index CK</u> |  |
| --- | --- | --- | --- | --- |
|  | OR (95% CI) | P | OR (95% CI) | P |
| **Statin type** |  |  |  |  |
| Atorvastatin | 1.49 (1.11, 1.99) | 0.007 | 1.35 (1.01, 1.81) | 0.04 |
| Lovastatin [Reference] | 1 |  | 1 |  |
| Pravastatin | 1.72 (1.00, 2.98) | 0.05 | 2.18 (1.39, 3.42) | 0.0007 |
| Simvastatin | 1.10 (0.93, 1.31) | 0.27 | 1.24 (1.01, 1.51) | 0.04 |
| **Statin daily dose** (irrespective of type) |  |  |  |  |
| <20 mg [Reference] | 1 |  | 1 |  |
| 20-40 mg | 1.11 (0.96, 1.29) | 0.14 | 1.38 (1.17, 1.63) | 0.0001 |
| >40 to <80 mg | 1.22 (0.90, 1.64) | 0.19 | 1.47 (1.17, 1.83) | 0.0007 |
| ≥80 mg | 2.92 (0.62, 13.78) | 0.18 | 1.97 (0.17, 22.29) | 0.58 |
| **Comorbidities** |  |  |  |  |
| Hypothyroidism | 22.13 (13.17, 37.19) | <0.0001 | 15.66 (9.13, 26.89) | <0.0001 |
| Hypertension | 1.37 (1.14, 1.64) | 0.0006 | 1.26 (1.04, 1.53) | 0.02 |
| Gout | 1.34 (1.09, 1.65) | 0.005 | 1.33 (1.05, 1.69) | 0.02 |
| Diabetes | 1.05 (0.91, 1.21) | 0.48 | 0.99 (0.84, 1.16) | 0.88 |
| Opioid abuse | 1.98 (1.30, 3.01) | 0.002 | 2.96 (1.73, 5.08) | <0.0001 |
| Dialysis | 1.90 (1.53, 2.35) | <0.0001 | 2.61 (2.01, 3.40) | <0.0001 |

Lastly, based on our algorithm development, we tested for relationships between comoribidities associated with SIM from the literature. Of these we found a statistically significant increases in SIM risk in statin users with hypothyroidism, hypertension, gout, opioid abuse and dialysis, for which hypothyroidism had the most dramatic increase (OR 15.65, 95% CI 9.12-26.83, P<0.0001, **Table 4**).

Finally, although new-onset diabetes is a common adverse effect of statin use, there was no relationship between diabetes diagnosis and SIM (OR 1.05, 95% CI 0.91-1.21, P=0.48).

## DISCUSSION

In this 3-part study, we developed an algorithm of statin-induced CK elevation, measured its performance through a variety of metrics, and applied it to our dataset to reveal clinical characteristics of SIM. To our knowledge, this is the largest and most racially-diverse study of its kind. Our resulting algorithm has applicability to a wide range of SIM studies from observational datasets and is easy to apply in high-throughput.

Existing algorithms designed to identify SIM in electronic health records are derived from study populations that are smaller or more homogenous than that of the current study. For example, Sai et al generated an algorithm from 5,109 statin users of a Japanese hospital^13^. Likewise, the Chan et al algorithms were derived from electronic health records of ∼1,200 SIM cases from a Singapore hospital^12^: this population was diverse (57% Chines, 17% Malay, 13% Indian, 13% Other), but not representative of a typical United States health system. In contrast, our study population was 55% white, 12% Black, 11% Hispanic, and 14% Asian by self-identified race/ethnicity. Although the algorithm steps are not impacted by population demographics, algorithm performance metrics may vary by population as response is known to differ by group^20^. Thus, the results of our study may be more generizable to North American populations compared to that of other publications.

Identifying clinical conditions associated with all-cause CK elevation was a critical step for the development of our algorithm. Unsurprisingly, most of the conditions that we had prespecified to interrogate had strong associations for this phenotype. Myocardial infarction, acute kidney failure, and strenuous exercise were among the predictors of all-cause CK elevation with the strongest associations regardless of whether participants were statin users or not. These results are consistent with previous literature ^21,22^ and provided a long list of factors for our algorithm to help us rule out secondary causes of CK elevation in statin users during identification of true SIM cases.

The performance of our full algorithm (applying steps 1 and 2) varied based on our manual chart review definition of a gold standard statin myopathy case. Among the randomly selected set of all-cause CK elevation cases, our algorithm was most sensitive (77%) for detecting statin-induced CK elevation when our gold standard definition was set as “certain” only (compared to certain+probable and certain+probable+possible). This indicates that the algorithm is best at detecting statin-induced CK elevation cases that are more clear cut (e.g., certain). The specificity of the algorithm was best as the gold standard became more inclusive (e.g., for certain+probable+possible gold standard cases, algorithm specificity was 85%). This suggests that the algorithm performs well at differentiating completely unlikely from all potential true cases. The sensitivity for the algorithm in detecting certain+probable+possible cases was 58%. This is a similar performance as an algorithm developed by Chai et al that had a sensitivity of 56%^13^, but a lower performance metric value than the best performing algorithm by Chan et al (72%-78% sensitivity)^12^. However, the Chan et al algorithm required keyword searching in the clinical notes, which may not be feasible or high-throughput in many datasets derived from electronic health records. Altogether, these findings have implications in choosing which situations are most suitable for the algorithm. Our algorithm would perform well for studies of large sample size where it’s more important to “rule out” CK elevation cases unrelated to statins than to “rule in” the most certain statin-induced CK elevation cases (e.g., a SIM phenotype for a large genome wide association study). For smaller studies, where it’s important to include only the most certain cases (e.g., sequencing the genome from a handful of statin users to identify rare, high penatrant variants associated with SIM), our algorithm may be a good starting point, but may need to be performed in conjunction with another tool to have a more confident set of certain cases.

Following algorithm validation, we conducted analyses in our algorithm-predicted statin-induced CK elevation cases to characterize this newly derived phenotype. We found a strong dose response for the odds of statin-induced CK elevation with higher doses expectedly linked to higher risk. When comparing our statin dosing variables, daily milligram strength had a stronger association with statin-induced CK elevation compared to daily LDL-C lowering intensity. These results are consistent with clinical trial data showing milligram dose, but not LDL-C lowering efficacy, to be associated with myopathy prevalence^23^.

Moreover, we observed that susceptibility to statin-induced CK elevation varied by statin type. Compared to our lovastatin reference, use of atorvastatin, pravastatin, and simvastatin were each associated with increased risk of SIM. Our data also showed that pravastatin was the statin type with the strongest risk for statin-induced CK elevation after adjusting for dose and other factors. It is commonly thought among clinical circles that hydrophilic statins (e.g., pravastatin, rosuvastatin) are less likely to cause SIM compared to those that are lipophilic (e.g., atorvastatin, simvastatin) due to an assumed reduction in skeletal muscle cell exposure. However, data including that of the current analysis does not strongly corroborate this belief. For example, clincal trial data showed that the incidence of statin-induced CK elevation with rosuvastatin 80 milligrams daily was greater than 80 milligrams per day of atorvastatin or simvastatin (and contributed to rosuvastatin 80 milligrams daily never being approved)^23^. An observational study with a similar design as the current report also does not show a strong correlation between lipophilicity and risk for SIM^24^. Lipophilic statins are more prone to drug-drug interactions^25^ and are more strongly incluenced by genetic polymorphisms in pharmacogenes^26^, both of which could increase drugs levels and the risk of SIM. These factors may be confounding associations between lipophilicity and SIM observed anecdotally. Among all comorbidities tested, hypothyroidism was by far the strongest predictor of statin-induced CK elevation. Prior studies demonstrate an association between SIM and hypothyroidism, whereas a link connecting myopathy with isolated hypothyroidism has not been reported^27^. With a 16-fold increased risk for statin-induced CK-elevation in the presence of hypothyroidism (no other comorbidity had >3-fold odds), our findings underscore the importance of monitoring thyroid levels in clinical settings prior to statin initiation and periodically thereafter. Altogether, the characterization of our algorithm-derived statin-induced CK elevation phenotype provides additional confirmation for the accuracy and future utility of our algorithm by replicating well-established SIM findings.

## CONCLUSION

In summary, we have developed a high throughput algorithm for detecting SIM in electronic health records and validated its accuracy using manual chart review. This methodological tool can enhance the quality of statin phenotypic data for large pharmacogenomic and pharmacoepidemiologic studies, results of which could inform health care providers in better optimizing statin therapy regimens to avoid adverse patient outcomes.

## Data Availability

All data produced in the present study are available upon reasonable request to the authors.

## Acknowledgements

We thank Payvand Milani, David Kirakossian and Albert Ma for their assistance in chart review.

## Author contributions

Conceptualization: AO, MWM, CI

Methodology: AO, MWM, CI, ML

Investigation: AO, ML

Funding acquisition: MWM, CI

Project administration: MWM, CI

Supervision: CI

Writing – original draft: AO, MWM

Writing – reviewing & editing: AO, MWM, CI, ML, RK, JP

## REFERENCES

1. Baigent, C. et al. Efficacy and safety of cholesterol-lowering treatment: prospective meta-analysis of data from 90,056 participants in 14 randomised trials of statins. Lancet 366, 1267–1278 (2005).

2. Grundy, S. M. et al. 2018 AHA/ACC/AACVPR/AAPA/ABC/ACPM/ADA/AGS/APhA/ASPC/NLA/PCNA guideline on the management of blood cholesterol: A report of the American College of Cardiology/American Heart Association Task Force on Clinical Practice Guidelines. Circulation 139, e1082–e1143 (2019).

3. Pencina, M. J. et al. Application of new cholesterol guidelines to a population-based sample. N. Engl. J. Med. 370, 1422–1431 (2014).

4. >Gomez Sandoval, Y.-H., Braganza, M. V. & Daskalopoulou, S. S. Statin discontinuation in high-risk patients: a systematic review of the evidence. Curr. Pharm. Des. 17, 3669–3689 (2011).

5. Mann, D. M., Woodward, M., Muntner, P., Falzon, L. & Kronish, I. Predictors of nonadherence to statins: a systematic review and meta-analysis. Ann. Pharmacother. 44, 1410–1421 (2010).

6. Zhang, H., Plutzky, J. & Turchin, A. Discontinuation of statins in routine care settings. Ann. Intern. Med. 159, 75–76 (2013).

7. El-Salem, K. et al. Prevalence and risk factors of muscle complications secondary to statins. Muscle Nerve 44, 877–881 (2011).

8. Cohen, J. D., Brinton, E. A., Ito, M. K. & Jacobson, T. A. Understanding Statin Use in America and Gaps in Patient Education (USAGE): an internet-based survey of 10,138 current and former statin users. J. Clin. Lipidol. 6, 208–215 (2012).

9. Buettner, C. et al. Statin use and musculoskeletal pain among adults with and without arthritis. Am. J. Med. 125, 176–182 (2012).

10. Alfirevic, A. et al. Phenotype standardization for statin-induced myotoxicity. Clin. Pharmacol.Ther. 96, 470–476 (2014).

11. Chapman, W. W. et al. Overcoming barriers to NLP for clinical text: the role of shared tasks and the need for additional creative solutions. J. Am. Med. Inform. Assoc. 18, 540–543 (2011).

12. Chan, S. L. et al. Development and validation of algorithms for the detection of statin myopathy signals from electronic medical records. Clin. Pharmacol. Ther. 101, 667–674 (2017).

13. Sai, K. et al. Development of a detection algorithm for statin-induced myopathy using electronic medical records. J. Clin. Pharm. Ther. 38, 230–235 (2013).

14. Rawson, E. S., Clarkson, P. M. & Tarnopolsky, M. A. Perspectives on exertional rhabdomyolysis. Sports Med. 47, 33–49 (2017).

15. Torres, P. A., Helmstetter, J. A., Kaye, A. M. & Kaye, A. D. Rhabdomyolysis: pathogenesis, diagnosis, and treatment. Ochsner J. 15, 58–69 (2015).

16. Leverenz, D., Zaha, O., Crofford, L. J. & Chung, C. P. Causes of creatine kinase levels greater than 1000 IU/L in patients referred to rheumatology. Clin. Rheumatol. 35, 1541–1547 (2016).

17. Oudman, I. et al. Creatine kinase is associated with failure of hypertension treatment. J. Hypertens. 31, 1025–1031 (2013).

18. Oni-Orisan, A. et al. Characterization of Statin Low-Density Lipoprotein Cholesterol Dose-Response Using Electronic Health Records in a Large Population-Based Cohort. Circ. Genom. Precis. Med. 11, e002043 (2018).

19. Ganna, A. et al. Risk prediction measures for case-cohort and nested case-control designs: an application to cardiovascular disease. Am. J. Epidemiol. 175, 715–724 (2012).

20. Liao, J. K. Safety and efficacy of statins in Asians. Am. J. Cardiol. 99, 410–414 (2007).

21. George, M. D., McGill, N.-K. & Baker, J. F. Creatine kinase in the U.S. population: Impact of demographics, comorbidities, and body composition on the normal range. Medicine (Baltimore) 95, e4344 (2016).

22. Biccard, B. M. Investigation of predictors of increased creatine kinase levels following vascular surgery and the association with peri-operative statin therapy. Cardiovasc. J. Afr. 20, 187–191 (2009).

23. Jacobson, T. A. Statin safety: lessons from new drug applications for marketed statins. Am. J. Cardiol. 97, 44C–51C (2006).

24. van Staa, T. P., Carr, D. F., O’Meara, H., McCann, G. & Pirmohamed, M. Predictors and outcomes of increases in creatine phosphokinase concentrations or rhabdomyolysis risk during statin treatment. Br. J. Clin. Pharmacol. 78, 649–659 (2014).

25. Bitzur, R., Cohen, H., Kamari, Y. & Harats, D. Intolerance to statins: mechanisms and management. Diabetes Care 36 Suppl 2, S325–30 (2013).

26. Voora, D. et al. The SLCO1B1*5 genetic variant is associated with statin-induced side effects. J. Am. Coll. Cardiol. 54, 1609–1616 (2009).

27. Bar, S. L., Holmes, D. T. & Frohlich, J. Asymptomatic hypothyroidism and statin-induced myopathy. Can. Fam. Physician 53, 428–431 (2007).

