## Supplementary Information for "Development and application of an algorithm for statin-induced myopathy based on electronic health record-derived structured elements"

**Supplementary Table 1.** All ICD and CPT codes used.

| | | ICD-9 | ICD-9 Procedure Codes | ICD-10-CM | ICD-10 Procedure Codes (PCS) | CPT4 |
|---|---|---|---|---|---|---|
| 01 | MI/STEMI/NSTEMI | 410 | | I21, I22 | | |
| 02 | Non-MI ACS | 411, 413 | | I20, I25, I25 | | |
| 03 | CABG/PCI-Stent (RP) | | 36.x, V45.82 | Z95.5, Z98.61 | 02120Z9, 021009W 0270346 02703ZZ 02703DZ 02CO3ZZ | 33510, 33511, 33512, 33513, 33514, 33515, 33516, 33517, 33518, 33519, 33521, 33522, 33523, 33530, 33533, 33534, 33535, 33536 92980, 92981, 92982, 92984, 92995, 92996, 92975, 92977 |
| 04 | Operations on valves and septa of the heart | | 35.0 | Q21.0 Q21.1 | | 33365, 33366, 33390, 33391,33404, 33405, 33406, 33410, 33411, 33413, 33414, 33415, 33416, 33417, 33420, 33425, 33427, 33430, 33460,33463, 33464, 33465, 33468, 33470, 33471, 33475, 33476, 33478, 33496, 33542, 33545, 33548, 33600, 33602, 33608, 33610, 33611, 33612, |

| | | | | | | |
|---|---|---|---|---|---|---|
| | | | | | | 33615, 33617, 33619, 33641, 33645, 33647, 33660, 33665, 33670, 33675, 33676, 33677, 33681, 33684, 33688, 33692, 33697, 33702, 33710, 33720, 33722, 33732, 33735, 33736, 33737, 33770, 33774, 33776, 33780, 33782, 33783, 33786, 33813, 33814, 33920 |
| 05 | Cardiogenic shock | 785.51 | | R57.0 | | |
| 06 | Hip replacement | | 81.51, 81.52 | | 0SR9*, 0SRA*, 0SRB*, 0SRE*, 0SRR*, 0SRS* | 27125, 27130, 27132, 27134, 27137, 27138, 27236 |
| 07 | Abdominal AAA repair | 38.34, 38.44, 38.64 | | | 04500ZZ 04503ZZ 04504ZZ 04B00ZZ 04B03ZZ 04B04ZZ 04R007Z 04R00JZ 04R00KZ 04R047Z 04R04JZ 04R04KZ 04U03JZ 04U04JZ 04V03DZ 04V04DZ | 33875, 33877, 34830, 34831, 34832, 35081, 35082, 35091, 35092, 35102, 35103 |
| 08 | Carotid endarterectomy | | 38.1 | | I03CK0ZZ | 35301, 35390 |

| | | | | | | |
|---|---|---|---|---|---|---|
| | | | | | 03CL0ZZ | |
| 10 | Knee arthroplasty/knee replacement | | 81.22, 81.54 | | 0SRC*, 0SRD*, 0SRT*, 0SRU*, 0SRV*, 0SRW* | 27438, 27440, 27441, 27442, 27443, 27445, 27446, 27447, 27486, 27487 |
| 11 | Spinal fusion | | 81.0 | | ORG*** OSG*** OSR0-8*** | 22532, 22533, 22548, 22551, 22554, 22556, 22558, 22586, 22590, 22595, 22600, 22610, 22612, 22630, 22633, 22800, 22802, 22804, 22808, 22810, 22812, 22880, 0195T, 0196T |
| 12 | Bladder cystectomy | | 57.1, 57.6 | | **0TSB0ZZ** | 47480, 47562, 47563, 47564, 47570, 47600, 47605, 47610, 47612, 47620, 47720, 47721, 47740, 47741 |
| 13 | Mastectomy | | 85.x | | **0HBV0ZZ** 0HTT0ZZ 0HTU022 | 19120, 19125, 19126, 19300, 19301, 19302, 19303, 19304, 19305, 19306, 19307, 19316, 19318, 19324, 19325, 19328, 19330, 19340, 19342, 19350, 19355, 19357, 19361, 19364, 19366, 19368, 19371, 19380 |

| | | | | | | |
|---|---|---|---|---|---|---|
| 14 | Hysterectomy | | 68.3, 68.4, 68.5, 68.6, 68.7, 68.8, 68.9 | | 0UT90ZZ<br>0UTC0ZZ | 58150, 58152, 58180, 58290, 58210, 58240, 58541, 58542, 58543, 58544, 58548, 58570, 58571, 58572, 58573, 58951, 58953, 58954, 58956, 59525, 58575 |
| 15 | Oophorectomy | | 65.2, 65.3, 65.4, 65.5, 65.6 | | 0UT14ZZ<br>0UT04ZZ | 58660, 58661, 58662, 58679, 58720, 58740, 58800, 58805, 58820, 58822, 58825, 58900, 58920, 58925, 58940, 58943, 58950, 58952, 58970 |
| 16 | Nephrectomy/ kidney transplant | | 55.4, 55.5, 55.6 | | 0TT0***<br>0TT1***<br>0TT2*** | 50220, 50225, 50230, 50234, 50236, 50240, 50250, 50280, 50290, 50320, 50400, 50405, 50441, 50442, 50443, 50445, 50446, 50447, 50448, 50449 |
| 17 | Heart transplant | | 37.5 | | **02YA*** ** | 33945, 33927, 33928 |
| 18 | Lobectomy/lung transplant | | 32.3, 32.4, 32.5, 32.6, 33.5 | | 0BBC***<br>0BBD***<br>0BBF***<br>0BBG***<br>0BBH***<br>0BBJ***<br>0BBK***<br>0BBL*** | 32440, 32442, 32445, 32480, 32482, 32484, 32486, 32488, 32491, 32501, 32503, 32504, 32505, 32506, 32507 |

| | | | | | | |
|---|---|---|---|---|---|---|
| | | | | | 0BBM***<br>OBYM*** | |
| 19 | Hepatectomy/Liver transplant | | 50.3, 50.4,<br>50.5 | | 0FT0***<br>0FY0*** | 47120, 47122,<br>47125, 47130,<br>47140, 47141,<br>47142, 47135, 47399 |
| 20 | Bone marrow transplant | | 41.0 | | 30230G0<br>30233G0<br>30240G0<br>30243G0<br>330250G0<br>30253G0<br>30260G0<br>30263G0<br>41.02<br>41.03<br>30230G1<br>30230G2<br>30230G3<br>30230G4<br>30233G1<br>30233G2<br>30233G3<br>30233G4<br>30240G1<br>30240G2<br>30240G3<br>30240G4<br>30243G1<br>30243G2<br>30243G3<br>30243G4<br>30250G1<br>30253G1<br>30260G1<br>30263G1 | 38204, 38220,<br>38242, 38232 |
| 21 | Pancreatectomy/pancreas transplant | | 52.5, 52.6,<br>52.7, 52.8,<br>52.9 | | 0FBG***<br>0FYG*** | 48550-56 |

| | | | | | | |
|---|---|---|---|---|---|---|
| 22 | Appendix surgery | | 47.x | | 0DBJ*** | 44900, 44950,44955, 44960, 44970, 44970 |
| 23 | Gastrectomy | | 43.5, 43.6, 43.7, 43.8, 43.9 | | 0DB7*** | 43610, 43611, 43620, 43621, 43622, 43631, 43632, 43633, 43634 |
| 24 | Pacemaker insertion | | 37.6, 37.7, 37.8, 37.9 | | 0JH636Z<br>02H63JZ<br>02HK3JZ<br>3E0132A | 33217, 33218, 33220, 33221, 33222, 33223, 33224, 33225, 33226, 33227, 33228, 33229, 33230, 33231, 33233, 33234, 33235, 33236, 33236, 33237, 33238, 33240, 33244, 33249, 33262, 33263, 33264, 33270, 33271, 33272, 33273 |
| 25 | Thyroid and parathyroid surgery | | 06 | | 0GTG***<br>0GTH***<br>0GTK***<br>0GTL***<br>0GTM***<br>0GTN***<br>0GTP***<br>0GTQ***<br>0GTR*** | 60000, 60200, 60210, 60212, 60220, 60225, 60240, 60252, 60254, 60260, 60270, 60271, 60280, 60281, 60500, 60502, 60505, 60512 |
| 26 | Splenectomy | | 41.43, 41.5 | | 07BP*** | 38100, 38101, 38102, 38115, 38120, 38200 |
| 27 | Cholecystectomy | | 51.2 | | 0FB4*** | 47562, 47563, 47564, 47570, 47600, 47605, 47610, 47612, |

| | | | | | | |
|---|---|---|---|---|---|---|
| | | | | | | 47620, 47621,<br>47640, 47641 |
| 28 | Resection/ removal of small or large intestine / colostomy | | 45.5, 45.6,<br>45.7, 45.8,<br>45.9 | | 0DBE***<br>0DBF***<br>0DBG***<br>0DBH***<br>0DBK***<br>0DBL***<br>0DBM***<br>0DBN***<br>0DBP*** | 44025, 44110,<br>44111, 44130,<br>44137, 44139,<br>44140, 44141,<br>44143, 44144,<br>44145, 44146,<br>44147, 44150,<br>44151, 44155,<br>44156, 44157,<br>44158, 44160,<br>44188, 44204,<br>44205, 44206,<br>44207, 44208,<br>44210, 44211,<br>44212, 44213,<br>44227, 44320,<br>44322, 44340,<br>44345, 44346,<br>44605, 44620,<br>44625, 44626, 57307 |
| 29 | Limb amputation | | 84.0, 84.1,<br>84.2 | | 0Y6**** | 23900, 23920,<br>24900, 24920,<br>24930, 24931,<br>25900, 25905,<br>25909, 25920,<br>25922, 25924,<br>25927, 25929,<br>25931, 26551,<br>26910, 26951,<br>26952, 27290,<br>27291, 27590,<br>27591, 27592,<br>27598, 27880,<br>27882, 27884,<br>27886, 27888,<br>27889, 28800,<br>28805, 28810,<br>28820, 28825 |

| | | | | | | |
|---|---|---|---|---|---|---|
| 30 | Acute kidney failure, dialysis | 584.9, V56 | N17.0-N17.2 N17.8-N17.9 | | | 90935, 90936, 90937, 90938, 90939, 90940, 90941, 90942, 90943, 90944, 90945, 90946, 90947, 90951, 90952, 90953, 90954, 90955, 90956, 90957, 90958, 90959, 90960, 90961, 90962, 90963, 90964, 90965, 90966, 90967, 90968, 90969, 90970, 99070, 99289, 99290, 99512, 99559 |
| 31 | Accidents/injuries | E800-E848, E900-E909, E910-E915, E916-E926, E928-E929, E980-E989 | | V00-Y99 excluding W00-19 | | |
| 32 | Falls | E880-E888 | | W00-19, Z91.81 | | |
| 34 | Sepsis / septic shock | 038 | | A41.2, A41.01, A41.02, A41.11, A41.4, A41.50, A41.3, A41.51, A41.52, A41.53, A41.59, A41.89, A47.27, A41.9 R65.20-R65.21 | | |
| 35 | Coma | 780.01 | | R40.20 | | |
| 36 | Ischemic Stroke | 433.01, 433.11, 433.21, 433.31, 433.81, 433.91, 434.01, 434.11, 434.91, 436.x | | I63.x | | |

| | | | | |
|---|---|---|---|---|
| 37 | Hemorrhagic stroke | 430.x, 431.x, 432.1, 432.9 | | I60.x, I61.x, I62.x |
| 38 | Lymphedema | 457.1 | | I89.0 |
| 39 | Convulsions (febrile, other) | 780.31, 780.39 | | R56.00, R56.9 |
| 40 | Convulsions (epilepsy) | 345.10-345.91 | | G40.309, G40.401, G40.409, G40.311, G40.319, G40.411, G40.419, G40.A01, G40.A09, G40.A11, G40.A19, G40.301, G40.201, G40.209, G40.211, G40.219, G40.001, G40.009, G40.101, G40.109, G40.011, G40.019, G40.111, G40.119, G40.821, G40.822, G40.823, G40.824, G40.501, G40.509, G40.801, G40.802, G40.811, G40.812, G40.089, G40.B01, |

| | | | | |
|---|---|---|---|---|
| | | | | G40.B09, G40.803, G40.804, G40.813, G40.814, G40.89, G40.B11, G40.B19, G40.901, G40.911, G40.919 |
| 41 | Paraplegia | 344.1 | | G04.1, G82.20, G82.21, G82.22 |
| 42 | Dehydration | 276.5, 276.0, 276.1 | | E87.0, E87.1, E87.6, E86.0 |
| 43 | HIV/AIDS | 042, V08 | | B20, Z21 |
| 44 | Burns | 941.3x, 941.4x, 941.5x, 942.3x, 942.4x, 942.5x, 943.3x, 943.4x, 943.5x, 944.3x, 944.4x, 944.5x, 945.3x, 945.4x, 945.5x | | T30-T32 |
| 45 | Diabetic ketoacidosis | 250.1 | | E11.69, E13.10, E10.10, E11.65, E11.69, E10.65 |
| 46 | Hypothyroidism | 243, 244.x | | E00.0, E00.1, E00.2, E00.9, E03.0, E03.1, E89.0, E03.2, E01.8, E03.3, E03.8, E03.9 |
| 47 | Pneumonia | 480.xx-486.xx | | J120, J121, J122, J1281, J123, J1289, J129, J13, J181, J150, J151, J14, J154, J153, J1520, J15211, J15212, J1529, |

| | | | | |
|---|---|---|---|---|
| | | | | J158, J155, J156, A481, J158, J159, J157, J160, J168, B250, A3701, A3711, A3781, A3791, A221, B440, J17, B7781, J180, J188, J189 |
| 48 | Heat stroke | 992.0 | | T67.0XXA |
| 49 | Overexertion from strenuous movement or load | E927.x | | X50.0 |
| 50 | Hypertension | 401,402,403,404,405 | | I10, I11, I12, I13, I14, I15, I16 |
| 51 | Gout | 274 | | M10 |
| 52 | Diabetes | | | |
| 53 | Opioid | | | |
| 54 | Skeletal Disease | '359.1', '359.2', '359.3', '359.5', '359.6', '359.7' | | 'G71.0', 'G71.1', 'G72.3', 'G73.7', 'G72.41' |
| 55 | Toxic myopathy | 359.4 | | G72.0, G72.2, T46.6 |
| 55 | Rhabdomyolysis | 728.88 | | M62.82 |

**Supplementary Table 2.** Criteria used to ascertain SAMS case status during manual chart review

| |
|---|
| **Certain (any of the following)** |
| Positive rechallenge (CK <4x after statin cessation and >4x upon rechallenge) |
| Definitive diagnosis by clinician |
| **Probable (does not meet Certain plus the following)** |
| Exclusion of other potential factors for CK >4x but no rechallenge or definitive diagnosis |
| **Possible (does not meet Certain or Probable plus any of the following)** |
| Mention of statin along with other potential factors together in the differential diagnosis |
| No mention of statin in notes, but statin is discontinued followed by ≥1 CKs <4x within a reasonable time period |
| Differential diagnosis not recorded or unclear, but statin discontinued followed ≥1 CKs <4x within a reasonable time period that cannot be linked to resolution of an acute disorder |
| **Unlikely (does not meet Certain, Probable, or Possible plus any of the following)** |
| Clinicians note one or more culprits of CK elevation that they believe are more likely than a statin |
| No temporal association between statin and CK elevation (e.g., patient hadn't been taking statin) |
| Differential diagnosis not recorded or unclear, but 1) statin is continued or 2) statin is discontinued plus resolution of an acute disorder followed by ≥1 CKs <4x within a reasonable time period |

**Supplementary Table 3.** Statin daily dose of the statin users with elevated CK (cases) and statin users with normal CK (control) at the time of statin initiation and at the time of elevated CK (case) or index CK (control).

| | | At First Statin Prescription | | | At Time of Elevated CK | | |
|---|---|---|---|---|---|---|---|
| | | **Elevated CK CASES** Statin users with CK ≥ 4 ULN† n= 5,430 | **CONTROLS** Statin Users with CK always < 2 ULN n=13,171 | **P** | **Elevated CK CASES** Statin users with CK ≥ 4 ULN† n= 5,430 | **CONTROLS** Statin Users with CK always < 2 ULN n=13,171 | **P** |
| **Atorvastatin** | <20 mg | 96 (1.8%) | 201 (1.5%) | 0.23 | 217 (4.0%) | 395 (3.0%) | 0.0005 |
| | 20-40 mg | 80 (1.5%) | 157 (1.2%) | 0.12 | 274 (5.0%) | 391 (3.0%) | <0.0001 |
| | 40-80 mg | 60 (1.1%) | 102 (0.8%) | 0.03 | 334 (6.2%) | 566 (4.3%) | <0.0001 |
| | ≥80 mg | 48 (0.9%) | 92 (0.7%) | 0.18 | 81 (1.5%) | 172 (1.3%) | 0.32 |
| **Lovastatin** | <20 mg | 1,273 (23.4%) | 3,303 (25.1%) | 0.02 | 775 (14.3%) | 2,295 (17.4%) | <0.0001 |
| | 20-40 mg | 1,452 (26.7%) | 3,602 (27.3%) | 0.40 | 910 (16.8%) | 2,592 (19.7%) | <0.0001 |
| | 40-80 mg | 1,150 (21.2%) | 2,572 (19.5%) | 0.01 | 808 (14.9%) | 2,047 (15.5%) | 0.26 |
| | ≥80 mg | 25 (0.5%) | 34 (0.3%) | 0.03 | 52 (1.0%) | 104 (0.8%) | 0.25 |
| **Pravastatin** | <20 mg | 9 (0.2%) | 14 (0.1%) | 0.29 | 34 (0.6%) | 74 (0.6%) | 0.60 |
| | 20-40 mg | 29 (0.5%) | 83 (0.6%) | 0.44 | 64 (1.2%) | 109 (0.8%) | 0.02 |
| | 40-80 mg | 26 (0.5%) | 61 (0.5%) | 0.89 | 65 (1.2%) | 128 (1.0%) | 0.17 |
| | ≥80 mg | 1 (0.0%) | 9 (0.1%) | 0.18 | 4 (0.1%) | 18 (0.1%) | 0.26 |
| **Rosuvastatin** | <20 mg | 2 (0.0%) | 3 (0.0%) | 0.60 | 4 (0.1%) | 8 (0.1%) | 0.75 |
| | 20-40 mg | 2 (0.0%) | 2 (0.0%) | 0.36 | 5 (0.1%) | 5 (0.0%) | 0.15 |
| | 40-80 mg | 2 (0.0%) | 3 (0.0%) | 0.60 | 5 (0.1%) | 9 (0.1%) | 0.59 |
| | ≥80 mg | NA | NA | NA | 1 (0.0%) | 5 (0.0%) | 0.50 |
| **Simvastatin** | <20 mg | 85 (1.6%) | 228 (1.7%) | 0.42 | 435 (8.0%) | 977 (7.4%) | 0.16 |
| | 20-40 mg | 192 (3.5%) | 543 (4.1%) | 0.06 | 502 (9.2%) | 1,290 (9.8%) | 0.25 |
| | 40-80 mg | 658 (12.1%) | 1,660 (12.6%) | 0.36 | 684 (12.6%) | 1,648 (12.5%) | 0.87 |
| | ≥80 mg | 240 (4.4%) | 502 (3.8%) | 0.05 | 176 (3.2%) | 338 (2.6%) | 0.01 |

**Supplementary Table 4.** Characteristics and Distribution of Acute[1] and Chronic Conditions[2] among Statin Users and Statin Non-Users at CK elevation

| | **Statin Users** | | | **Statin Non-Users** | | |
|---|---|---|---|---|---|---|
| | **Elevated CK CASES** Statin users with CK ≥ 4x ULN† n= 5,430 | **CONTROLS** Statin Users with CK always < 2x ULN n=13,171 | **P** | **Elevated CK CASES** Statin non-users with CK ≥ x4 ULN† n= 5,726 | **CONTROLS_1¶** Statin non-users with CK always < 2x ULN n=22,904 | **P*** |
| **Acute Conditions, n (%)** | | | | | | |
| MI/STEMI/NSTEMI/RP | 1,399 (25.76%) | 23 (0.17%) | <0.0001 | 1,467 (25.62%) | 27 (0.12%) | <0.0001 |
| Non-MI ACS | 771 (14.20%) | 57 (0.43%) | <0.0001 | 841 (14.69%) | 87 (0.38%) | <0.0001 |
| Major surgery | 138 (2.54%) | 13 (0.10%) | <0.0001 | 147 (2.57%) | 14 (0.06%) | <0.0001 |
| Acute kidney failure | 1,114 (20.52%) | 19 (0.14%) | <0.0001 | 1,125 (19.65%) | 24 (0.10%) | <0.0001 |
| Accidents/ Injuries | 318 (5.86%) | 11 (0.08%) | <0.0001 | 371 (6.48%) | 21 (0.09%) | <0.0001 |
| Falls | 630 (11.60%) | 16 (0.12%) | <0.0001 | 679 (11.86%) | 26 (0.11%) | <0.0001 |
| Sepsis / septic shock | 757 (13.94%) | 7 (0.05%) | <0.0001 | 814 (14.22%) | 8 (0.03%) | <0.0001 |
| Stroke | 290 (5.34%) | 13 (0.10%) | <0.0001 | 310 (5.41%) | 17 (0.07%) | <0.0001 |
| Convulsions (febrile, epilepsy, other) | 166 (3.06%) | 10 (0.08%) | <0.0001 | 170 (2.97%) | 12 (0.05%) | <0.0001 |
| Dehydration | 730 (13.44%) | 9 (0.07%) | <0.0001 | 813 (14.20%) | 13 (0.06%) | <0.0001 |
| Pneumonia | 717 (13.20%) | 16 (0.12%) | <0.0001 | 755 (13.19%) | 18 (0.08%) | <0.0001 |
| Overexertion from strenuous movement or load | 10 (0.18%) | 1 (0.01%) | <0.0001 | 10 (0.17%) | 1 (0.00%) | <0.0001 |
| SBP > 180 mmHg and/or DBP> 120 mmHg | 597 (10.99%) | 26 (0.20%) | <0.0001 | 665 (11.61%) | 39 (0.17%) | <0.0001 |
| Uric acid > 7.2 mg/dL in men or > 6.5 mg/dL in women | 197 (3.63%) | 23 (0.17%) | <0.0001 | 207 (3.62%) | 30 (0.13%) | <0.0001 |
| HbA1C > 9% | 108 (1.99%) | 31 (0.24%) | <0.0001 | 113 (1.97%) | 48 (0.21%) | <0.0001 |
| TSH > 3 mIU/L | 387 (7.13%) | 81 (0.61%) | <0.0001 | 408 (7.13%) | 110 (0.48%) | <0.0001 |
| **Chronic Conditions** | | | | | | |
| Hypothyroidism | 756 (13.92%) | 96 (0.73%) | <0.0001 | 801 (13.99%) | 135 (0.59%) | <0.0001 |
| Hypertension | 4,815 (88.67%) | 10,582 (80.34%) | <0.0001 | 5,084 (88.79%) | 18,331 (80.03%) | <0.0001 |
| Gout | 738 (13.59%) | 1,172 (8.90%) | <0.0001 | 793 (13.85%) | 2,087 (9.11%) | <0.0001 |
| Diabetes | 1,714 (31.57%) | 3,639 (27.63%) | <0.0001 | 1,824 (31.85%) | 6,319 (27.59%) | <0.0001 |
| Opioid abuse | 216 (3.98%) | 121 (0.92%) | <0.0001 | 227 (3.96%) | 333 (1.45%) | <0.0001 |
| Skeletal Disease | 5 (0.09%) | 5 (0.04%) | 0.1477 | 5 (0.09%) | 11 (0.05%) | 0.26 |
| HIV/AIDS | 29 (0.53%) | 4 (0.03%) | <0.0001 | 30 (0.52%) | 6 (0.03%) | <0.0001 |
| Dialysis | 1,160 (21.36%) | 676 (5.13%) | <0.0001 | 1,204 (21.03%) | 1,614 (7.05%) | <0.0001 |
| **Clinical Diagnosis** | | | | | | |
| Toxic Myopathy[3] | 36 (0.66%) | 1 (0.01%) | <0.0001 | 40 (0.70%) | 7 (0.03%) | <0.0001 |
| Rabdomyolysis[4] | 1,698 (31.27%) | 12 (0.09%) | <0.0001 | 1,816 (31.71%) | 24 (0.10%) | <0.0001 |
| Toxic Myopathy or Rabdomyolysis 2 weeks after CK elevation | 1,293 (23.81%) | 0 (0.00%) | <0.0001 | 1,379 (24.08%) | 0 (0.00%) | <0.0001 |

1: (acute conditions) 7 days prior or 3 days later to elevated CK 4 x ULN in cases or 7 days prior or 3 days later to index CK elevation date of controls

2: (chronic conditions) Between 5 years prior to first statin prescription up to elevated CK or index CK elevation date

*t-test or Chi-Square test

† and the CK value is within the statin prescription window or no more than 2 weeks past the end of the prescription window

¶ matched to cases 1:4 on age, sex, race and ensuring that the control is alive and a KP member at the index CK elevation date and that the statin initiation in controls is within +/- 365 days of the statin initiation of the corresponding case

**single measure preceding closest in time.

3: ICD-9 code 359.4 or ICD-10 codes G72.0, G72.2 or T46.6, ascertained between first statin prescription and two weeks after elevated CK in cases or index CK elevation date for controls.

4: ICD-9 code 728.88 or ICD-10 code M62.82, ascertained between first statin prescription and two weeks after elevated CK in cases or index CK elevation date for controls.

**Supplementary Table 5.** Multivariate Conditional Logistic Regression Predicting Elevated CK≥ 4 ULN among Statin Users without consideration of metabolic factors (n=5,340 cases with CK ≥ 4 ULN; n=13,171 controls with CK always < 2 ULN.)

| | | OR (95% CI) | P |
|---|---|---|---|
| **Statin Type and Daily Dose at the Time of CK Elevation** | | | |
| Atorvastatin | <20 mg | 1.40 (0.96, 2.03) | 0.08 |
| | 20-40 mg | 1.80 (1.26, 2.59) | 0.001 |
| | 40-80 mg | 1.58 (1.12, 2.22) | 0.01 |
| | ≥80 mg | 1.45 (0.86, 2.46) | 0.17 |
| Lovastatin | <20 mg | 0.92 (0.75, 1.13) | 0.43 |
| | 20-40 mg | 1.00 [Ref] | |
| | 40-80 mg | 1.22 (1.00, 1.49) | 0.05 |
| | ≥80 mg | 1.38 (0.73, 2.60) | 0.32 |
| Pravastatin | <20 mg | 2.16 (0.99, 4.68) | 0.05 |
| | 20-40 mg | 3.06 (1.67, 5.60) | 0.0003 |
| | 40-80 mg | 2.14 (1.16, 3.93) | 0.01 |
| | ≥80 mg | 1.30 (0.20, 8.57) | 0.79 |
| Rosuvastatin | <20 mg | 3.97 (0.30, 53.15) | 0.30 |
| | 20-40 mg | 0.46 (0.01, 18.65) | 0.68 |
| | 40-80 mg | 0.95 (0.10, 9.46) | 0.97 |
| | ≥80 mg | 0.83 (0.00, 437.77) | 0.95 |
| Simvastatin | <20 mg | 1.48 (1.14, 1.92) | 0.003 |
| | 20-40 mg | 1.28 (1.00, 1.63) | 0.05 |
| | 40-80 mg | 1.48 (1.19, 1.84) | 0.0004 |
| | ≥80 mg | 1.91 (1.38, 2.66) | 0.0001 |
| **Acute Conditions** | | | |
| MI/STEMI/NSTEMI/RP | | 199.67 (97.13, 410.47) | <0.0001 |
| Non-MI ACS | | 10.22 (5.84, 17.86) | <0.0001 |
| Major surgery | | 19.57 (8.36, 45.79) | <0.0001 |
| Acute kidney failure, dialysis | | 67.94 (30.38, 151.94) | <0.0001 |
| Accidents/ Injuries | | 61.11 (18.21, 205.06) | <0.0001 |

| | | |
|---|---|---|
| Falls | 105.99 (41.59, 270.09) | <0.0001 |
| Sepsis / septic shock | 23.48 (8.94, 61.66) | <0.0001 |
| Stroke | 16.15 (6.51, 40.08) | <0.0001 |
| Convulsions (febrile, epilepsy other) | 6.33 (2.84, 14.13) | <0.0001 |
| Dehydration | 83.49 (30.22, 230.68) | <0.0001 |
| Pneumonia | 31.95 (14.69, 69.51) | <0.0001 |
| Overexertion from strenuous movement or load | 19.09 (0.96, 379.80) | 0.05 |
| SBP > 180 mmHg and/or DBP> 120 mmHg | 17.94 (9.54, 33.77) | <0.0001 |
| Uric acid > 7.2 mg/dL in men or >6.5 mg/dL in women | 18.87 (9.44, 37.75) | <0.0001 |
| HbA1C > 9% | 5.76 (3.12, 10.65) | <0.0001 |
| TSH > 3 mIU/L | 10.95 (7.25, 16.56) | <0.0001 |
| **Chronic Conditions** | | |
| Hypothyroidism | 10.42 (6.82, 15.91) | <0.0001 |
| Hypertension | 1.19 (1.02, 1.38) | 0.03 |
| Gout | 1.24 (1.01, 1.51) | 0.04 |
| Diabetes | 1.03 (0.91, 1.17) | 0.64 |
| Opioid abuse | 2.88 (1.82, 4.58) | <0.0001 |
| Skeletal Disease | 0.63 (0.02, 23.75) | 0.80 |
| HIV/AIDS | 44.72 (6.14, 325.82) | 0.0002 |
| Dialysis | 2.02 (1.61, 2.52) | <0.0001 |

**Supplementary Table 6.** Performance of algorithms for SIM detection based on manual chart review

| Certain cases only | Algorithm defined cases | Positive Chart review cases | Negative chart review cases | Sensitivity | Specificity | PPV | NPV | F |
|---|---|---|---|---|---|---|---|---|
| **All-Cause CK** | Positive | 21 | 74 | 100% | 0% | 22% | 0% | 0.36 |
| | Negative | 0 | 0 | | | | | |
| **Step 1 alone** | Positive | 20 | 42 | 95% | 43% | 32% | 97% | 0.48 |
| | Negative | 1 | 32 | | | | | |
| **Step 1 + 2** | Positive | 16 | 17 | 76% | 77% | 48% | 92% | 0.59 |
| | Negative | 5 | 57 | | | | | |

| Certain + Probable | Algorithm defined cases | Positive Chart review cases | Negative chart review cases | Sensitivity | Specificity | PPV | NPV | F |
|---|---|---|---|---|---|---|---|---|
| **All-Cause CK** | Positive | 29 | 66 | 100% | 0% | 31% | 0% | 0.47 |
| | Negative | 0 | 0 | | | | | |
| **Step 1 alone** | Positive | 28 | 34 | 97% | 48% | 45% | 97% | 0.61 |
| | Negative | 1 | 32 | | | | | |
| **Step 1 + 2** | Positive | 19 | 14 | 66% | 79% | 58% | 84% | 0.62 |
| | Negative | 10 | 52 | | | | | |

| Certain + Probable + Possible | Algorithm defined cases | Positive Chart review cases | Negative chart review cases | Sensitivity | Specificity | PPV | NPV | F |
|---|---|---|---|---|---|---|---|---|
| **All-Cause CK** | Positive | 43 | 52 | 100% | 0% | 45% | 0% | 0.62 |
| | Negative | 0 | 0 | | | | | |
| **Step 1 alone** | Positive | 40 | 22 | 93% | 58% | 65% | 91% | 0.77 |
| | Negative | 3 | 30 | | | | | |
| **Step 1 + 2** | Positive | 25 | 8 | 58% | 85% | 76% | 71% | 0.66 |
| | Negative | 18 | 44 | | | | | |

| Key | Algorithm defined cases | Positive Chart review cases | Negative chart review cases | Sensitivity | Specificity | PPV | NPV | F |
|---|---|---|---|---|---|---|---|---|

| | | | | | | | | |
|---|---|---|---|---|---|---|---|---|
| | Positive<br>Negative | True Positive<br>False Negative | False Positive<br>True Negative | TP/(TP+FN) | TN/(TN+FP) | TP/(TP+FP) | TN/(TN+FN) | (2xPPV x Sensitivity)/(PPV + Sensitivity) |

**Supplementary Table 7.** Independent associations of statin type, statin daily dose (irrespective of type) and comorbidities with SIM case/control status.

| | At time of elevated/index CK | |
|---|---|---|
| | **OR (95% CI)** | **P** |
| **Statin Type** | | |
| Atorvastatin | 1.27 (0.95, 1.68) | 0.11 |
| Lovastatin [Reference] | 1 | |
| Pravastatin | 2.11 (1.34, 3.31) | 0.001 |
| Simvastatin | 1.18 (0.97, 1.44) | 0.09 |
| **Z score of statin daily dose** (irrespective of type) | 1.15 (1.07, 1.25) | 0.0004 |
| **Comorbidities*** | | |
| Hypothyroidism | 15.64 (9.12, 26.82) | <0.0001 |
| Hypertension | 1.26 (1.04, 1.53) | 0.02 |
| Gout | 1.33 (1.05, 1.69) | 0.02 |
| Diabetes | 0.99 (0.84, 1.16) | 0.92 |
| Opioid abuse | 2.96 (1.72, 5.08) | <0.0001 |
| Dialysis | 2.64 (2.03, 3.44) | <0.0001 |

**Supplementary Table 8.** Association of trajectories of statin daily dose (irrespective of type) between initial daily dose vs. daily dose at time of elevated CK with SIM or index CK with controls

| | SIM Cases<br>n= 1,257 | Controls<br>n=3,047 | OR (95% CI) |
|---|---|---|---|
| **No change** | 621 (49.4%) | 1,668 (54.7%) | 1.00 [REF] |
| **Increased dose** | 543 (43.2%) | 1,165 (38.2%) | 1.22 (1.04, 1.43) |
| **Decreased dose** | 93 (7.4%) | 214 (7.0%) | 0.93 (0.70, 1.25) |
